# Completeness and timeliness of a contact tracing intervention during the COVID-19 pandemic: an outcome evaluation of the California Contact Tracing Support Initiative

**DOI:** 10.64898/2026.02.05.26345670

**Authors:** Soim Park, Kyra H. Grantz, Kyu Han Lee, Erica N. Rosser, Lana Feras Aldos, Riaa Dutta, Laura-Marie Peeples, Emily S. Gurley, Melissa A. Marx, Elizabeth C. Lee

## Abstract

**Objectives:** To describe COVID-19 case investigation and contact tracing outcomes from an intervention conducted in two California counties under the California Contact Tracing Support Initiative (CCTSI).

**Methods:** We analyzed contact tracing program metrics in two counties using de-identified individual-level records from COVID-19 cases and their contacts. Interviews with 47 program staff were conducted and analyzed using thematic analysis.

**Results:** In one county, 64% of assigned COVID-19 cases (2,722 of 4,279) were successfully interviewed, with 77% (2,105) of interviews occurring within 48 hours of receiving test results; 63% of assigned contacts (418 of 660) were notified, with 82% (348) of notifications occurring within 48 hours of contact elicitation. In a second county, 52% of cases (1,743 of 3,352) were interviewed, with 48% (1,621) interviewed occurring within 48 hours; 79% of contacts (155 of 196) were notified, with 82% (127) notified within 48 hours. CCTSI staff reported faster receipt of test results in the early months and positive perceptions of wrap-around service referrals.

**Conclusions:** The performance of the CCTSI contact tracing intervention differed across two California counties. In both counties, there was moderate to high case interview completion and contact notification overall and within 48 hours of case and contact identification, but relatively few contacts were elicited per case. The perceived benefit of several program features was high among staff and stakeholders. The evaluation highlighted generalizable challenges in contact tracing investigations and linking contact and case records in contact tracing databases.

## Introduction

Following the first confirmed case in the United States (US) on January 20, 2020,^1^ coronavirus disease 2019 (COVID-19) had caused roughly 385,000 deaths nationwide by December 2020.^1^ Case investigation and contact tracing were among the principal strategies implemented for disease control before a vaccine was introduced in late 2020.^2^ Local health jurisdictions (LHJs) like county health departments were primarily responsible for community contact tracing with some support from state health departments, while institutions (e.g., schools, prisons) also sometimes performed contact tracing in their specialized populations.^3,4^

Case investigation typically involves contacting a confirmed case of infectious disease, eliciting information on individuals in close contact with that case (i.e., contacts) during the timeframe when the case may have been infectious, and advising isolation.^5,6^ Contact tracing involves notifying the contacts of their potential exposure and encouraging quarantine. The joint process of case investigation and contact tracing (heretofore, “contact tracing”) is important for liaising and disseminating information and resources to the community, and across the highly localized public health authorities, there was substantial heterogeneity in innovations, implementations, and challenges among contact tracing programs. Lack of trust in public health authorities or the technology companies that developed digital data collection tools and concerns about misinformation and misuse of private information were oft-cited barriers to contact tracing.^7–9^ Reaching marginalized communities was also a major challenge of contact tracing programs, exacerbating health inequalities.^10,11^ Black and Hispanic populations experienced higher rates of COVID-19 incidence, hospitalization and mortality than white populations.^12,13^ At the same time, minority and immigrant communities expressed heightened concerns about participating in contact tracing activities due to a complex interplay of historical, cultural and legal factors that engendered mistrust of government authorities.^14,15^

To mitigate these barriers and support California state’s contact tracing activities, the Public Health Institute (PHI), a public health implementation organization, and Kaiser Permanente (KP), a large healthcare provider, launched the California Contact Tracing Support Initiative (CCTSI). Between December 2020 and July 2021, the program supported contact tracing for members of the KP healthcare system who tested positive for COVID-19 (cases) and their community contacts in seven California counties. Our team conducted a comprehensive evaluation of this contact tracing intervention as implemented in two California counties, Fresno and San Bernardino, and this article reports on program outcomes. The primary evaluation goals were to describe program metrics related to contact tracing completeness and timeliness and perceptions related to providing wrap-around service referrals to program beneficiaries. Secondarily, we compared the performance of the contact tracing activities between the CCTSI and the five California LHJs.

## Methods

### Study design and setting

Our study evaluated CCTSI activities conducted in Fresno County from December 2020 to July 2021 and San Bernardino County from February to July 2021. The intervention was triggered when a positive COVID-19 test result for a KP healthcare network member was imported into a CCTSI database developed for contact tracing data management. The intervention sought to build trust with program beneficiaries by highlighting the case investigator’s affiliation with KP during contact tracing phone calls and deploying investigators trained in empathetic engagement and whose native languages and cultural backgrounds matched those of beneficiaries. These staff were arranged into “micro-teams” comprising investigators who called cases or contacts, resource coordinators who referred beneficiaries to “wrap-around services” available at community, county, state, and national levels (e.g., COVID-19 test scheduling, hygiene kits, face masks, food, housing, or utilities assistance) that could facilitate isolation and quarantine, and supervisors who provided oversight. Micro-team members managed and coordinated contact tracing data collection using the same CCTSI database receiving test results.

### Quantitative data collection and analysis

We analyzed de-identified individual-level records of COVID-19 cases from the CCTSI counties and their community contacts from the CCTSI database. The number and types of wrap-around service referral requests were also extracted. Data were cleaned in consultation with CCTSI epidemiologists familiar with the database structure. We also requested monthly aggregate contact tracing metrics from California LHJs from December 2020 through July 2021 (**Supplementary Table 1**) to provide baseline context on community-wide contact tracing performance during the program period.

In brief, positive test results were *received* in the CCTSI database and cases were *assigned* to investigators. Investigators *attempted* outreach a maximum of four times, and those successfully *reached* were subsequently *interviewed* (italicized labels described further in **Figure 1)**. If interviewed cases reported close contact with individuals, investigators *elicited* their names and contact details, contacts were *assigned* to investigators, and outreach was *attempted*. Contacts that completed interviews were considered successfully *notified* about their potential exposure to COVID-19. The evaluation impact model provides detail on how program inputs and activities are linked conceptually to desired program outputs and outcomes (**Supplementary Figure 1**).^16^

**Figure 1.**
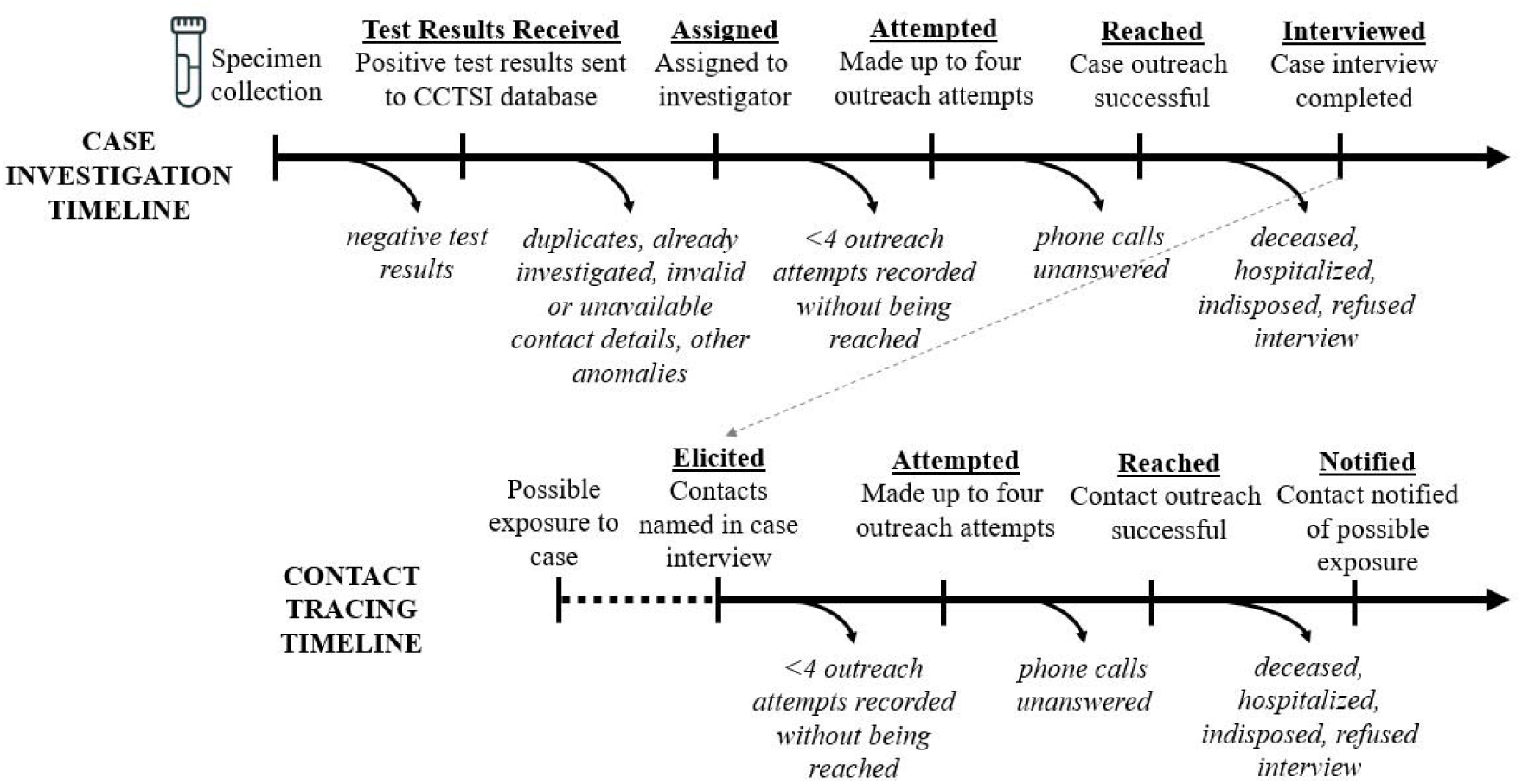
Conceptual diagram depicting case investigation and contact tracing activities. Case investigation (top row) and contact notification (bottom row) status definitions are provided under each label. Reasons for potential loss of cases and contacts throughout an investigation are provided in italics under the outflow arrows. The grey dashed arrow indicates that contacts are elicited during case interviews.

Completeness was assessed by measuring percent of assigned, attempted, reached, and interviewed test-positive cases, percent of contacts assigned to investigation among those elicited, and percent of attempted, reached, and notified contacts among those assigned to investigation (**Figure 1**).^17^ Timeliness was measured as percent of cases and contacts with attempted outreach or successful interview within 24 and 48 hours after receipt of test results or contact elicitation.^17^ Timeliness in test results reporting was measured by percent of cases reported within 24 and 48 hours of specimen collection. Timeliness metrics for LHJs were reported within 1 and 2 calendar days of specimen collection, providing coarser temporal information than the hourly metrics. Metrics for wrap-around service referral requests included percent of requests created, requests closed, and types of services requested. Completeness, timeliness, and wrap-around service referral requests were described cumulatively and monthly across the CCTSI counties and compared descriptively to the available LHJ metrics and time periods.

### Qualitative data collection and analysis

To understand the program’s purpose, intended design, implementation, and the experiences of staff and partners, we conducted in-depth interviews with stakeholders including high and mid-level management and micro-team members. To capture relevant inputs from the stakeholders, we iteratively refined interview guides as other study results became available (e.g., quantitative, document review, other qualitative interviews) (See **Supplementary Files 1-2** for final interview guides). Initial interview participants were selected by reviewing roles and responsibilities identified in program documents and through initial discussions with PHI and KP. PHI and KP leadership supported recruitment by notifying potential participants via staff meetings and email that our evaluation team might contact them for interviews. Snowball sampling was also deployed during interviews with participants suggesting additional interview candidates. Sample size was guided by the principle of data saturation, whereby interviews continued until no new significant themes or perspectives emerged. This article presents the subset of this data relevant to program implementation context and wraparound services.

Interviews were audio-recorded except in two cases in which recording consent was declined; for these two interviewees, the interviewer took detailed interview notes upon which the analysis was based. In addition, interview teams summarized the interview by writing their own notes immediately after each interview. All recorded interviews were transcribed verbatim and uploaded with field notes into NVivo version 14 (QSR International, Melbourne, Australia). A team of four coders completed an inductive open-coding exercise to generate codes for thematic analysis. After adding five deductive codes adopted from the impact model, the team generated 70 initial codes which were refined into a final codebook with 32 codes for coding transcripts.

### Ethical approval

The Johns Hopkins Bloomberg School of Public Health Institutional Review Board granted an exemption for the analysis of quantitative secondary data from CCTSI and LHJs and approved qualitative data collection and analysis (IRB No. 17623). All qualitative study participants provided oral informed consent and could opt in to receive $25 gift cards as a token of appreciation.

## Results

From the CCTSI database, we obtained records for 4,529 test-positive KP members at the Fresno site and 3,352 from the San Bernardino site. Monthly and cumulative contact tracing metrics were obtained from five California LHJs. Across program roles, we contacted 100 individuals and 47 agreed to be interviewed. Qualitative interviews were conducted with 24 stakeholders and 23 micro-team members (**Supplementary File 3).**

### Program beneficiary characteristics

There were 3,140 total interviewed cases and traced contacts associated with the Fresno site and 1,898 at the San Bernardino site (**Table 1**). Approximately 40% of cases and contacts in both counties were younger than 35 years old, and more than half (54% in Fresno; and 52% in San Bernardino) were female. About 50% of cases and contacts were Hispanic, which resembles the Hispanic or Latino population proportion in the two counties (55%).^18^ At the San Bernardino site, 9% of cases and contacts were interviewed or notified in Spanish.

**Table 1.**
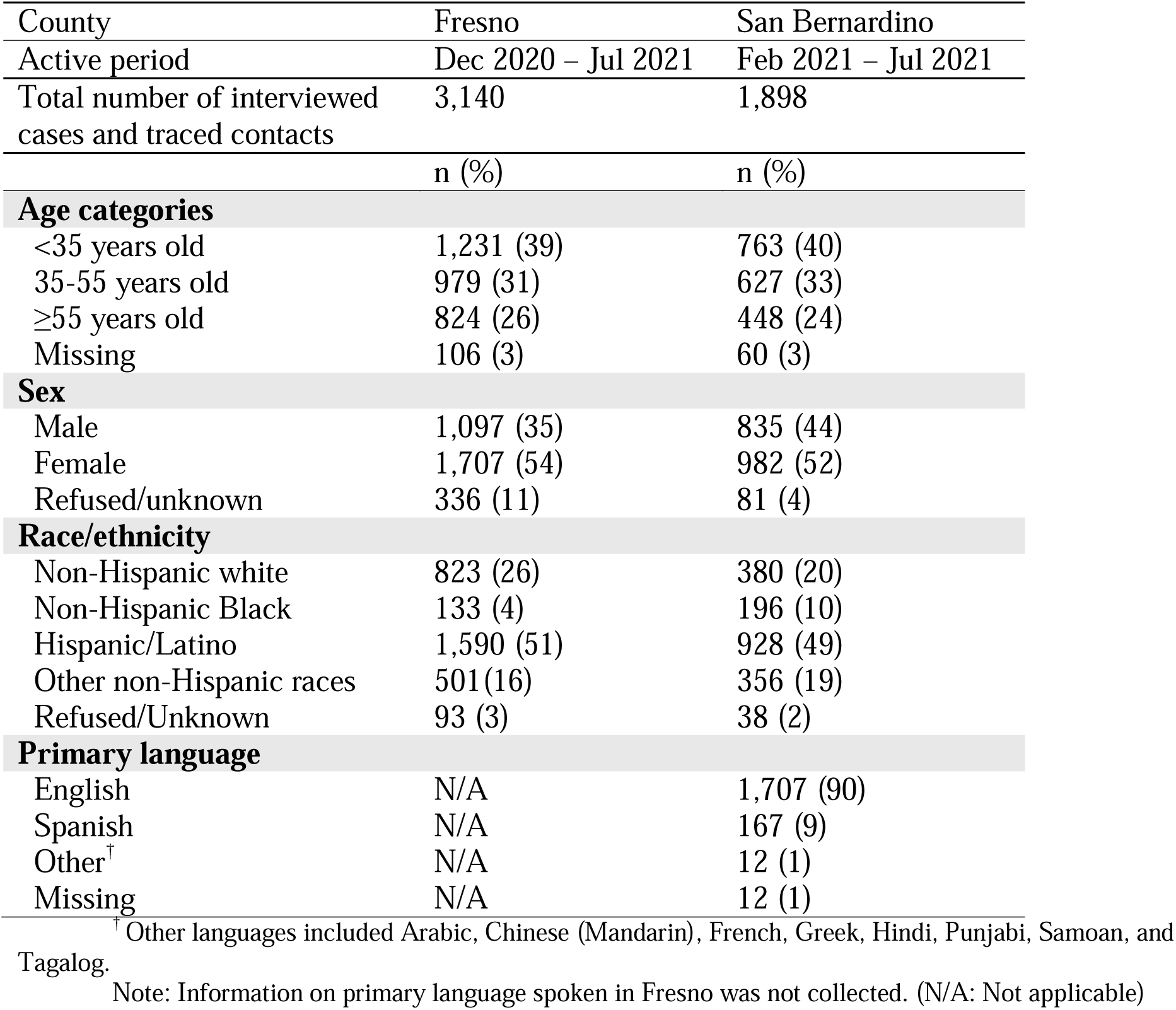
Sociodemographic information of interviewed cases and notified contacts (%) in CCTSI: Fresno and San Bernardino counties.

| County | Fresno | San Bernardino |
| --- | --- | --- |
| Active period | Dec 2020 – Jul 2021 | Feb 2021 – Jul 2021 |
| Total number of interviewed cases and traced contacts | 3,140 | 1,898 |
|  | n (%) | n (%) |
| **Age categories** |  |  |
| <35 years old | 1,231 (39) | 763 (40) |
| 35-55 years old | 979 (31) | 627 (33) |
| ≥55 years old | 824 (26) | 448 (24) |
| Missing | 106 (3) | 60 (3) |
| **Sex** |  |  |
| Male | 1,097 (35) | 835 (44) |
| Female | 1,707 (54) | 982 (52) |
| Refused/unknown | 336 (11) | 81 (4) |
| **Race/ethnicity** |  |  |
| Non-Hispanic white | 823 (26) | 380 (20) |
| Non-Hispanic Black | 133 (4) | 196 (10) |
| Hispanic/Latino | 1,590 (51) | 928 (49) |
| Other non-Hispanic races | 501 (16) | 356 (19) |
| Refused/Unknown | 93 (3) | 38 (2) |
| **Primary language** |  |  |
| English | N/A | 1,707 (90) |
| Spanish | N/A | 167 (9) |
| Other<sup>†</sup> | N/A | 12 (1) |
| Missing | N/A | 12 (1) |

<sup>†</sup> Other languages included Arabic, Chinese (Mandarin), French, Greek, Hindi, Punjabi, Samoan, and Tagalog.
Note: Information on primary language spoken in Fresno was not collected. (N/A: Not applicable)

### Assessing outcomes of the contact tracing activities

#### Completeness

Of 4,529 Fresno-site COVID-positive cases, 95% (n=4,279) were assigned to investigators (**Table 2**). Of assigned cases, 92% (n=3,923) had up to four call attempts, 66% (n=2,807) were reached by phone, and 64% (n=2,722) were successfully interviewed. All 3,352 San Bernardino-site COVID-positive cases were assigned to an investigator and 97% (n=3,243) had call attempts, 65% (n=2,193) were reached, and 52% (n=1,743) were successfully interviewed.

**Table 2.** Cumulative completeness of case investigation and contact tracing for CCTSI in Fresno and San Bernardino Counties.

| County | Fresno | San Bernardino |
| --- | --- | --- |
| Period of analysis | Dec 2020 – Jul 2021 | Feb 2021 – Jul 2021 |
| **Case investigations:** |  |  |
| Cases with positive test results, n | 4,529 | 3,352 |
| Assigned, n (% of imported) | 4,279 (95) | 3,352 (100) |
| Attempted, n (% of assigned) | 3,923 (92) | 3,243 (97) |
| Reached, n (% of assigned) | 2,807 (66) | 2,193 (65) |
| Interviewed, n (% of assigned) | 2,722 (64) | 1,743 (52) |
| Cases eliciting any contacts, n (% of interviewed) | 1,222 (45) | 1,352 (78) |
| **Contact notifications:** |  |  |
| Total contacts elicited, n | 4,277 | 4,943 |
| Assigned, n (% of total contacts elicited) | 660 (15) | 196 (4) |
| Attempted, n (% of assigned) | 587 (89) | 188 (96) |
| Reached, n (% of assigned) | 481 (73) | 163 (83) |
| Notified, n (% of assigned) | 418 (63) | 155 (79) |

Contacts were elicited from 45% (n=1,222) of cases interviewed in Fresno and 78% (n=1,352) of cases interviewed in San Bernardino (**Table 2**). A total of 4,277 and 4,943 contacts were elicited during case investigations in Fresno and San Bernardino Counties respectively, but only contacts with a valid name and phone number were assigned out for investigation – 660 contacts (15% of elicited contacts) in Fresno and 196 contacts (4% of elicited contacts) in San Bernardino. Of contacts assigned to investigation at the Fresno site, 89% (n=587) had call attempts, 73% (n=481) were reached by phone, and 63% (n=418) were successfully notified. Among assigned contacts in San Bernardino, 96% (n=188) had call attempts, 83% (n=163) were reached, and 79% (n=155) were successfully notified. Monthly completeness metrics are reported in **Supplementary Figures 2-5**.

Some cases and contacts were reached but neither interviewed nor notified. In Fresno, 85 cases were reached but not interviewed because they were deceased (n=24, 6% of 4,279 assigned cases), hospitalized or indisposed (n=33, 8%), or refused the interview (n=28, 7%). In San Bernardino, 450 cases were reached but not interviewed. The majority of these were refusals (n=356, 11% of 3,352 assigned cases), but some were deceased (n=14, <1%) or hospitalized or indisposed (n=79, 2%). All assigned contacts across both counties who were reached but not notified refused interviews (Fresno: n= 63, 10% of 660 assigned contacts; San Bernardino: n=8, 4% of 196 assigned contacts).

The CCTSI had attempted, reached, and interviewed case completeness metrics that fell within the range of reported LHJ metrics (**Supplementary Table 1**). Contact-related metrics were sparse and heterogeneous across LHJs; compared to available data, contact notification completeness achieved by CCTSI was in the higher end of the range reported by LHJs. The proportion of contacts elicited during case investigations that were converted into contact tracing investigations (i.e., assigned contacts) was lower for CCTSI (15% in Fresno and 4% in San Bernardino) compared to LHJs (20-80%).

CCTSI sought to improve completeness by leveraging the trust that call recipients might have in their healthcare provider. The initial Fresno site call script advised investigators to introduce themselves as calling on behalf of KP. However, investigators reported that the phone number from which they called was provided by a cloud-based communication system and did not match the Fresno County area code, thus leading call recipients to have suspicions about scam or spam calls. Concurrent public alerts about phone scams or phishing attacks related to COVID-19 created heightened awareness about call number issues among beneficiaries. As one supervisor reported,

> *“Whenever we would call, a 916-phone number would show up [to beneficiaries]. When we would call, they [beneficiaries] would say ‘why is it a 916 if you’re calling from Kaiser Fresno?’ During that period on the news, [there were warnings about] the scam callers.”*

Qualitative data suggested that beneficiaries became more receptive to investigator phone calls after CCTSI started sending pre-call text messages later in the program implementation. Interviews also indicated that case investigations and contact notifications for household members were sometimes performed on the same call. It is unclear whether and how frequently contemporaneous notifications were recorded in the database, but this may explain some of the loss of contacts between the *elicited* and *assigned* stages in the quantitative metrics.

#### Timeliness

At the Fresno site, most cases occurred in December 2020 (66%, 2,840 out of 4,279 assigned cases), coinciding with a winter COVID-19 surge (**Supplementary Figure 2**). At least one attempt was made to reach 4,005 cases in Fresno, and 72% (n=2,875) of them were reached within 24 hours of test result receipt (**Table 3**). The median time from test result receipt to first outreach was 4 hours (interquartile range (IQR) 2-26 hours). Among 4,279 assigned cases, 50% (n=2,105) were interviewed within 48 hours. The median time from test result receipt to case interview was 27 hours (IQR 5-77 hours) and 74% (n=429) of the 578 contacts with at least one outreach attempt were called within 24 hours. Seventy-one percent (n=296) of the 418 successfully notified contacts received their notification within 24 hours of contact elicitation.

**Table 3.** Cumulative timeliness of case investigation and contact tracing for CCTSI in Fresno and San Bernardino Counties. IQR: interquartile range.

| County | Fresno | San Bernardino |
| --- | --- | --- |
| Period of analysis | Dec 2020 – Jul 2021 | Feb 2021 – Jul 2021 |
|  | n (%) | n (%) |
| **Case investigations:** |  |  |
| **From specimen collection to receipt of test results** |  |  |
| N = Imported cases with specimen collection date | 4,368 | 3,159 |
| ≤24 hours (% of new cases imported) | 318 (7) | 0 (0) |
| >24, ≤48 hours (% of new cases imported) | 1,269 (29) | 799 (25) |
| **From receipt of test results to first outreach** |  |  |
| N = Cases with at least one timestamped attempt or were successfully reached | 4,005 | 3,228 |
| ≤24 hours (% with at least one attempt) | 2,875 (72) | 2,880 (89) |
| >24, ≤48 hours (% with at least one attempt) | 391 (10) | 223 (7) |
| Median time in hours (IQR) | 4 (2-26) | 3 (2-6) |
| **From receipt of test results to case interview** |  |  |
| N = Assigned cases | 4,279 | 3,352 |
| ≤24 hours (% of assigned) | 1,654 (39) | 1,244 (37) |
| >24, ≤48 hours (% of assigned) | 451 (11) | 377 (11) |
| Median time in hours (IQR) | 27 (5-77) | 8 (4-28) |
| **Contact notifications:** |  |  |
| Assigned contacts | 660 | 196 |
| **From contact elicitation to first outreach** |  |  |
| N = Contacts with at least one timestamped attempt or reached | 578 | 187 |
| ≤24 hours (% with at least one attempt) | 429 (74) | 134 (72) |
| >24, ≤48 hours (% with at least one attempt) | 59 (10) | 23 (12) |
| Median time in hours (IQR) | 2 (0.6-24) | 3 (0.5-25) |
| **From contact elicitation to contact notification** |  |  |
| N = Contacts notified | 418 | 155 |
| ≤24 hours (% of notified) | 296 (71) | 106 (68) |
| >24, ≤48 hours (% of notified) | 52 (12) | 21 (14) |
| **From case specimen collection to contact notification, Median time in days (IQR)** | 4 (3-6) | 4 (3-5) |
| **From receipt of case test results to contact notification, Median time in hours (IQR)** | 32 (7-98) | 28 (6-55) |

In San Bernardino, among 3,228 cases that were attempted at least once, 89% (n=2,880) were reached within 24 hours of test result receipt and the median time from case import to first outreach was 3 hours (IQR 4-28 hours) (**Table 3**). Among 3,352 assigned cases, 37% (n=1,244) and 11% (n=377) were interviewed in 24 hours and between 24 and 48 hours, respectively, and the median time from receiving test results to interviewing the case was 8 hours (IQR 4-28 hours). Most contacts (n=157, 84%) who had at least one outreach attempt were reached within 48 hours. Of 155 notified contacts, 68% (n=106) were notified within 24 hours.

There was substantial heterogeneity in the timeliness of LHJ activities. Across five LHJs, the percentage of cases successfully interviewed within 1 day of test result receipt varied from 12% to 73%, with a median of 42% (**Supplementary Table 2**). Only two LHJs were able to provide contact notification timeliness data, and the reported percentages of contacts notified within 1 day of contact elicitation were 50% and 37%. CCTSI was generally quicker than LHJs at completing case interviews and contact notifications (**Table 3**).

Multiple interview participants reported that the CCTSI database initially received test results more quickly than LHJ data systems. Thirty-six percent of Fresno cases (n=1,587 of 4,368 cases with specimen collection date) and 25% of San Bernardino cases (n=799 of 3,159) had test results within 48 hours of specimen collection (**Table 3**), while the average time from specimen collection to test result receipt at LHJs was 2 to 4.2 days (**Supplementary Table 2**). While LHJ data systems transferred COVID-19 test results across multiple databases, the CCTSI database was “*the first to get the [COVID-19] diagnosis*” more proximally from KP, thus reducing the time from specimen collection to case import. As one stakeholder stated, “*I think CCTSI held an advantage [in getting data faster than LHJs] for a long time. That advantage we held onto for a while meant people were getting notified sooner*.” As time went on, data import into California’s contact tracing database improved until it eventually matched and exceeded CCTSI data import speeds. We could not directly validate the data import timeliness findings because the LHJ metrics were reported in units of days and advantages were thought to be on the order of hours.

### Assessing effects of wrap-around service referrals

Of the interviewed cases and notified contacts, 43% in Fresno and 21% in San Bernardino requested wrap-around service referrals (**Supplementary Table 3**), which far exceeded the percentage of similar requests reported by LHJs (LHJ1 0%; LHJ2 2%; LHJ5 2%). Requests for food and utilities assistance were the most reported in both counties, though 44% (n=171) of San Bernardino requests recorded no specific service types. As resource coordinators served only to inform beneficiaries about available resources, data on whether beneficiaries used these resources was unavailable.

Resource coordinators worked together to curate and maintain lists of resources available for different populations and service types and responded to wrap-around service referral requests from beneficiaries. To verify that resources remained available, one resource coordinator described, “*I call around organizations depending on what they [beneficiaries] need, because I don’t wanna send people to dead ends*.” When there were investigator referrals, resource coordinators used their verified resource lists to link program beneficiaries with relevant organizations. Resource coordinators reported that during COVID-19 case surges, they prioritized responding to requests from marginalized individuals (e.g., families without food or water and experiencing homelessness). Resource coordinators were also highly sensitive to the plight of undocumented populations and perceived that wrap-around service referrals were particularly helpful when there was “*fear of utilizing any public resources*” due to the possibility of identity disclosure.

## Discussion

Our study evaluated CCTSI’s intervention in two California counties and is one of few that investigated the outcome of innovations in call-based contact tracing activities. The intervention sought to overcome barriers to high contact tracing completeness and timeliness by retrieving COVID-19 test results and case contact information through data systems from a large healthcare provider in California and deploying methods designed to enhance trust between investigators and beneficiaries. Our evaluation of CCTSI in two counties found that investigations were successfully completed for 64% of cases and 63% of contacts in Fresno and 52% of cases and 79% of contacts in San Bernardino. For each of the two counties, over one-third of case investigations (39% in Fresno and 37% in San Bernardino) were completed within 24 hours of test result receipt, and nearly three-fourths of contacts (74% in Fresno and 72% in San Bernardino) were notified within 24 hours of elicitation from a case investigation. Unlike other contact tracing programs where missing or incorrect beneficiary information was a significant barrier to completeness,^19^ the program had names and phone numbers for the vast majority of test-positive individuals because they were KP members. However, roughly one third of cases did not answer investigator phone calls after multiple attempts, indicating that supplemental outreach methods could still have been useful. Program staff reported that pre-call text messages enhanced call response rates, but this claim could not be investigated with quantitative program metrics, and the prevalence of contact tracing scam calls and messages could be a barrier to effective implementation of this approach in some settings.^20^

The process of eliciting contacts during case interviews and achieving contact notifications is a critical component of contact tracing, as quarantining contacts will have limited onward disease transmission.^21,22^ CCTSI faced significant challenges in converting elicited contacts into active contact tracing investigations, and we were unable to investigate this further in the program records as contact elicitation and contact investigation records could not be linked in the CCTSI database. Informed by our qualitative data collection, we hypothesize that irregularities in recordkeeping for household member contacts and missing contact phone numbers contributed to the apparent contact tracing challenges.^23^ Several other US studies reported low levels of contact elicitation and notification.^24–26^ One US-based study found that phone-based interviews elicited more contacts than automated text message surveys,^27^ while another reported that COVID-19 cases with select health and demographic characteristics were more likely to provide close contact information.^28^ The importance of building trust between investigator and cases to improve contact elicitation is well-documented,^29–31^ but a related sub-study from our CCTSI evaluation suggested that beneficiary concerns about privacy and stigma may have outweighed the perceived benefits of reporting contact details in this instance.^32^ Future work should seek to develop strategies to minimize barriers to contact elicitation and improve data linkage between case investigation and contact notification records to enable better monitoring and evaluation of the full contact tracing process.

Returning test results and completing contact tracing activities in a timely manner is critical to efficacy,^33,34^ as quarantine only prevents onward transmission if it begins during an exposed contact’s incubation period.^36,37^ Guidance from Resolve to Save Lives recommended benchmarks of ≤ 2 days for returning test results, ≤ 1 day for case investigation, and ≤ 1 day for contact tracing,^35^ but in reality, testing delays in cases posed a significant challenge.^1,24,36^ CCTSI sought to improve testing turnaround times by linking its database to alternate data feeds, namely KP electronic medical records. Roughly 70% of test results in the CCTSI database were received 2 days after specimen collection, suggesting upstream delays in laboratory testing, data transfer, or entry of test results outside of CCTSI’s control. However, once test results were received, activities proceeded relatively quickly compared to metrics reported by LHJs, with over one-third of cases interviewed within 24 hours (similar to the median value reported by LHJs), and nearly three-fourths of contacts notified within 24 hours of contact elicitation (faster than any LHJ providing data). Had there not been challenges in convincing beneficiaries that CCTSI was affiliated with their primary healthcare provider, which was the original program intent, it is possible that the “in-house” nature of the contact tracing would have enhanced trust between investigators and beneficiaries and increased timeliness, as was reported by other studies with similar program features.^37,38^

During the COVID-19 pandemic, concerns about childcare responsibilities and financial hardship were reported as barriers to following isolation and quarantine guidelines, particularly among already vulnerable populations.^39–41^ When individuals could be linked to wrap-around services such as medical, mental health, food, and other support, studies found that cases and contacts were more likely to isolate and quarantine.^42^ Likewise, CCTSI integrated wrap-around service referrals into their program model to enhance isolation or quarantine, and food and utility support referrals were most frequently requested by CCTSI beneficiaries.^40,41^ While our evaluation measured neither beneficiary utilization of referred services nor the impact of these referrals on isolation and quarantine, program staff reported positive perceptions of this aspect of the program in addressing urgent beneficiary needs.

Our evaluation faced several challenges that limit the interpretation of our results. While we reported that CCTSI conducted timelier investigations than LHJs, these metrics are not directly comparable. LHJs served the general population and performed outreach with a mix of text message surveys and phone calls, while CCTSI served only KP healthcare network members and their contacts via phone call outreach. Further timeliness metrics were reported in units of days for LHJs and hours for CCTSI, which made it challenging to validate qualitative data claims suggesting that CCTSI was able to receive test results more quickly than LHJs in the early program period. In addition, qualitative data suggested that CCTSI prioritized activities that would directly help beneficiaries – rapid notification, information dissemination, and wrap-around services — over clean recordkeeping. This, in addition to database design choices about how fields were linked, meant that metrics on outreach attempts, and assigned and notified contacts may be particularly underestimated in our study. Finally, our evaluation provided a limited view of program impact; we only collected qualitative data on the perceptions and experiences of providers (e.g., people who designed or directly carried out the program) and not program beneficiaries (i.e., cases or contacts who received contact tracing phone calls), and we did not estimate the program’s impact on reducing COVID-19 transmission in the community.

## Conclusion

The CCTSI program implemented a wide range of contact tracing activities and wrap-around service referrals across multiple counties in California, and the investigation completeness and timeliness achieved by the program was on par with or better than that observed in other US jurisdictions in the same period. Beyond acknowledging the enormous human resources and training effort it takes to roll out a successful contact tracing program, future efforts should learn from the outcomes and innovations of CCTSI, namely in reducing delays in laboratory testing turnaround time, setting clear definitions and database linkages for case contacts, and enhancing contact elicitation.

## Data Availability

All data produced in the present study are available upon reasonable request to the authors.

## Acknowledgments

Special thanks to CCTSI staff and stakeholders for sharing information and documents about the program, including Marta Induni, Diane Royal, and Dana Williamson, the CCTSI epidemiologists and data team for their support in accessing and interpreting the CCTSI program metrics, and program staff at the local health jurisdictions that shared contact tracing program metrics.

