## Supplementary File 1 for "Completeness and timeliness of a contact tracing intervention during the COVID-19 pandemic: an outcome evaluation of the California Contact Tracing Support Initiative"

**Supplementary File 1**. Interview guide used for stakeholders

**Program evaluation of the California Contact Tracing Support Initiative for COVID-19**

**Key Informant Interview Guide for Management and Leadership**

**OPENING**

Thank you for agreeing to speak with me today. The purpose of this interview is to learn more about the California Contact Tracing Support Initiative (CCTSI). I will ask you some questions and these questions do not have right or wrong answers. I am interested in how this program was developed, implemented, and integrated into communities. I am also interested in your opinion on the programs' impact and effectiveness.

**GENERAL QUESTIONS/CCTSI PURPOSE**

- Can you tell me about your role at work within PHI or KP? (e.g., division of organization, main responsibilities)
- How were you involved with the CCTSI (*Probe for whether their work is more in program design or implementation*)?
- Why did PHI/KP initiate CCTSI (i.e., for what purpose or at whose request)?

**AIM 1: PROGRAM DESIGN, PROCESS, FACILITATORS, BARRIERS, AND PROGRAM ROLLOUT**

I would like to talk to you more about your experience creating the CCTSI program.

**DESIGN**

- Can you please describe how this CCTSI program was originally designed?
- How were you involved in designing the program?
- What were the facilitators or barriers to conceptualizing the program design? How did you overcome barriers?
- What were the program activities?
- What was the level of community involvement during this stage?
- Can you describe what considerations were made when developing this program?
- Did you consider privacy issues? Stigma?
- How did any of those considerations factor into the design/approach?
- Did people receive two contact tracing calls? One from CCTSI and one from the health department?
- Did this process change at any point?
- Was it different in different locations i.e., Fresno vs. San Bernadino? Why?

**PILOT PROGRAM**

- What can you tell me about the pilot program that was implemented in San Jose by KP?
- How successful was the program? What were the shortcomings of the pilot?
- How did experiences from the pilot program fit into the CCTSI implemented by PHI?

**TERMINOLOGY/LANGUAGE**

- Several of the program documents mention hiring from and serving the communities most affected/impacted by COVID-19. How does the program define most affected/impacted communities?
- How did you arrive at this definition?
- Do you think the definition was uniformly understood by other program staff and stakeholders?
- What led you to target the communities you did?
- What does “clinically integrated” really mean? *How* was the program clinically integrated?
- How were the contact tracing and contact notification scripts developed?
- Who was responsible for/involved in determining the content of the scripts?
- How did the content of scripts change overtime?
- What was the result of any changes to the scripts?
- What considerations were made when developing the content of the scripts? Epidemiological? Sociocultural?
- How did you address the challenge of having micro team members follow a script with carrying on an unscripted conversation?
- Some of the language in the script was more suggestive than prescriptive i.e., “we recommend you isolate”. Was this intentional? Why? Did it change over time? *(Probe for if TH had the “authority” to use certain language i.e., were they deputized by the county health department to do contact tracing?)*

**IMPLEMENTATION**

- Can you please describe the initial rollout of the CCTSI program?
- Was the program implemented as intended?
- If not, how, and why did the program make the changes?
- What were the strategies you used to facilitate the implementation process?
- What were the barriers?
- How did you overcome barriers?
- Can you please identify factors that impacted the CCTSI program implementation?
- *Probe:* Did any material factors (e.g., resources, time) impact CCTSI program implementation?
- *Probe:* Did any work-environment factors (e.g., workload, interpersonal dynamics) impact CCTSI program implementation?
- *Probe:* Did any contextual factors (e.g., epidemiological changes, politics) impact CCTSI program implementation?
- The incorporation of cultural components, like cultural competency and linguistic diversity, into the program was a unique feature. Do you think this component has been employed appropriately/effectively?
- How would you describe the working dynamics between stakeholders, leadership, and implementors during the program rollout?
- What kind of staffing changes did you notice during your time with CCTSI?
- *Probe for indications of turnover, probe for changes at different levels i.e., CTer vs. Supervisor etc.*
- How effective were contracting and subcontracting arrangements that were established to support program implementation and evaluation?

**AIM 2: PERCEPTION OF CCTSI PROGRAM AND RECOMMENDED IMPROVEMENTS**

I would like to talk to you about how the program was implemented, if it met expectations, its strengths, and weaknesses, and how you would modify this program for future implementation.

- How would you describe your direct experience with the program?
- How do you think other program stakeholders/partners/coworkers/staff/ community members would describe their direct experience with the program?
- What is your opinion on the overall effectiveness of program rollout/implementation at the Fresno/Sonoma/San Bernardino sites?
- If there were any downside of the program, how would you modify this?
- What positive and negative effects have arisen from the program (*probe:* anticipated or not)?
- What do you think were the strengths and weaknesses of the program?
- What have been the critical success factors and barriers to achieving program goals?
- Knowing what you know now/having seen the program implemented if you could go back to the drawing board – what would you do differently to improve program design and implementation?
- What are overall recommendations for future implementation of this program?
- *(Follow up depending on informant)* What performance monitoring and quality improvement processes should exist in future implementations?
- Do you think the program met its goal of hiring from/serving communities most affected/impacted by COVID-19?

**ADDITIONAL QUESTIONS DEPENDING ON INFORMANT TYPE:**

- To what extent were community residents involved with the program?
- What roles were made available to residents?
- How did PHI and KP (Kaiser Permanente) recruit local residents to work for these sites?
- Can you tell me about an event where leadership and management incorporated a local hire’s feedback on the CCTSI program?
- Please describe the career development opportunities made for employees.
- Can you share how this program positively impacted the community?
- Please describe components of this programs hiring and career development that need improvement.
- What were remote interactions with community members like?
- Do you attribute any success or ease in rapport with community members/cases/contacts and micro-team members to their being from those same communities or their CCTSI training?

**CLOSING**

- We are almost done with our interview. Is there anything else you would like to share about the California Contact Tracing Support Initiative?
- Are there any other people either within or outside your organization with whom it would be helpful for me to speak and get ideas around CCTSI designing and implementation?

This is the end of our interview today. Thank you very much for participating in this interview. Do you have any questions for me? [Answer any questions]. If you have any additional thoughts or questions, feel free to contact me, the principal investigator - Dr. Melissa Marx, or the Johns Hopkins Institutional Review Board (IRB).

**Supplementary File 2**. Interview guide used for micro-team members (i.e., case investigators and resource coordinators)

**Program evaluation of the California Contact Tracing Support Initiative for COVID-19**

**In-depth Interview Guide for Fresno/San Bernardino Site Micro-teams**

**OPENING**

Thank you for agreeing to speak with me today. The purpose of this interview is to learn more about the California Contact Tracing Support Initiative (CCTSI). I will ask you a few questions and these questions do not have right or wrong answers. I am interested in how the CCTSI program was rolled out at the Fresno/Sanoma/San Bernardino site. I am also interested in how you think of the programs impact on the communities.

**BACKGROUND/OPENING cont.**

I would like to begin by asking you a few brief background questions.

- How would you describe your role on the CCTSI?
- Can you walk me through your typical day at work?
- What are your main responsibilities?
- How long have you been working on this team/serving this role?
- Are you originally from this community?
- Do you currently reside in this community?
- Tell me how you became involved with CCTSI.
- What was your motivation for applying for this job?
- Were there any professional reasons that influenced your involvement?
- Any expected job-related advantages or disadvantages?

**AIM 2: PERCEPTION OF CCTSI PROGRAM AND RECOMMENDED IMPROVEMENTS**

I would like to talk about your experiences with recruitment, onboarding, training, and working with this program. Please share your thoughts on how this program was designed, its impact, strengths, and weaknesses, as well as recommendations for future versions.

**IMPLEMENTATION**

Recruitment

- Can you please describe what the recruitment/hiring process was like for you?
- Was there anything that made that process of recruitment/hiring easy/difficult for you? How did you manage difficulties?
- Please describe ways you would modify this experience.

Onboarding

- Can you please describe what the onboarding process was like for you?
- Was there anything that made that process of onboarding easy/difficult for you? How did you manage difficulties?
- Please describe ways you would modify this experience.

Training

- Can you please describe what the training process was like for you?
- Was there anything that made that process training easy/difficult for you? How did you manage difficulties?
- Please describe ways you would modify this experience.
- Tell me about the different training modules you had to complete?
- How did these trainings prepare you for your role?
- Once you started working, did you think there was anything else that you should have been trained on to help you perform your job better?

Working

- After being hired and trained, you started working. What is your work with the organization like?
- How was the relationship/communication with leadership?
- How did this relationship/communication impact the program roll out?
- How was the relationship with coworkers (e.g., other contact tracers)?
- How was morale while you were working with CCTSI? Yours, others? Did this change over time?
- What are the factors that influenced your interaction with the local community?
- Do you think whether you are from the Fresno/Sanoma/San Bernardino community is an important factor to build rapport with the people you contact?
- What were remote interactions with community members like?
- How would you improve these interactions?
- In your opinion, how has this program impacted the local community?
- Once you started working, were there any changes made to the program? If so, what and do you know why? Tell me more about that process.
- *Note to interviewer: probe for different possible factors (e.g., material: resources, time, workload; cultural: work environment, interpersonal dynamics including interpersonal cultural dynamics; or contextual: epidemiological changes, politics) impact CCTSI program implementation. Also, probe for timeline changes e.g., did unstructured processes become more streamlined/clear overtime?*
- How do you think other program leadership/partners/coworkers/staff/community members would describe their direct experience with the program?
- Why do you think that is?
- What kind of staffing changes did you notice during your time with CCTSI?
- *Probe for indications of turnover, probe for changes at different levels i.e., CTer vs. Supervisor etc.*

Off-boarding

- Can you please describe what the off-boarding process was like for you?
- Was there anything that made that process of off-boarding easy/difficult for you? How did you manage difficulties?
- Please describe ways you would modify this experience.

**STRENGTHS AND WEAKNESSES**

- When you think of your personal experiences with CCTSI from the time you were hired to today, if you were a program designer and could modify the CCTSI to better help the people you contacted what kind of changes would you make?
- *[If the participant has suggestions,]* Have you shared those thoughts with anyone in CCTSI? *(Probe to see how they provide feedback i.e., the mechanism for doing so, if their feedback was taken seriously, if they felt comfortable/able to give feedback etc.)*
- What do you think are the strengths of this CCTSI program?
- What do you think are the weaknesses of this CCTSI program?

**AIM 3: HIRING AND WORKFORCE DEVELOPMENT**

I would like to talk about your experiences with hiring and workforce development in the CCTSI program.

- Did you have time to participate in the work force development activities?
- Were you interested in in the work force development activities?
- How did working in this program impact you personally?
- Has this position changed the way you think or feel about your career?
- Has working in this program influenced your future job seeking?
- How has the program supported you professionally?
- How well did you feel you were able to do your job?
- Can you share a time when you felt that you did your job well?
- How prepared did you feel for your specific role?
- What do you think played a part in making you feel more or less prepared to do your job well?
- What do you think could help you improve your performance?
- In your opinion what additional support should the program provide?
- To employees?
- To the community?
- Can you please share any recommendations you would make if the program were going to continue in the future or be rolled out somewhere new?

**ROLE SPECIFIC QUESTIONS**

I would now like to talk about your experience as a supervisor/resource coordinator/contact tracer.

- Supervisor/Manager (responsibilities include supervising staff during a pandemic, time sheet management, managing virtual teams)
- Was there a rapport within the members of the micro-team?
- Were there any challenges to being the liaison between program leadership and the rest of the micro-team?
- Resource Coordinator /Community Outreach Specialist (responsibilities include training on COS role, referral process, resources, and CBOs)
- Please share a time when you interacted with the local community?
- Did they seem impacted by this program?
- Please describe what factors you think made it easier/harder for this program to work with CBOs?
- What can you tell me about any scripts you used during your work as a RC/COS?
- How closely did you stick to the script during calls?
- Did you have to make any adaptations to the script? What? Why?
- Was there anything that made it easier or more difficult to follow during calls?
- What did you think of the wording that was used in the scripts?
- Contact Tracer/Contact Notifier
- In what ways does your background help you connect with the people you call? How did your training help?
- Was there a rapport between you and those you contacted?
- How did you create a rapport? What strategies did you use?
- What made that process easier or more difficult?
- Any barriers to contacting COVID-19 positive community residents and their contacts?
- Because your contact to COVID-19 positive community residents and their contacts may discuss their private information did you confront any privacy-related issues when you contacted these people?
- Did you hear any COVID-19 positive community residents, and their contacts express concern about being stigmatized because they had COVID-19 or had contact with positive patients?
- What did you think about the script you were assigned to use for calls?
- How closely did you stick to the script during calls?
- Did you have to make any adaptations to the script? What? Why?
- Was there anything that made it easier or more difficult to follow during calls?
- What did you think of the wording that was used in the scripts?
- How long did a contact tracing/notifying call typically last? How did clients/contacts react to the length of the call?
- Did your training address call length?
- How did you manage this?
- What system were you using to record data during your contact tracing/notification calls?
- What did you think about using MARI/CalConnect?
- Was there anything that made the process of using it during calls  easy/difficult for you? How did you manage difficulties?
- How well did your training prepare you for using MARI/CalConnect?
- What recommendations do you have for how MARI/ CalConnect could have been adapted to improve your working experience?
- Did you share these thoughts with any of your supervisors? If so, what happened next?
- Please describe ways you would modify this experience.

**CLOSING**

- We’re almost done with our interview. Is there anything else you would like to share about California Contact Tracing Support Initiative?
- Are there any other people either within or outside your organization with whom it would be beneficial for me to speak?

This is the end of our interview today. Thank you very much for participating in this interview. Do you have any questions for me? [Answer any questions]. If you have any additional thoughts or questions, feel free to contact me, the principal investigator Dr.  Melissa Marx, or the Johns Hopkins Institutional Review Board (IRB).

**Supplementary File 3**. Qualitative participants

More than half of the stakeholders we interviewed (54%) were affiliated with PHI (**Supplementary** **Table 1-1**). Among 23 MT members, we interviewed 12 case investigators (52%), four resource coordinators (17%), five supervisors (22%), and two team managers (9%). Although we tried to recruit relatively balanced number of MT members from both study sites, 18 (78%) MT members interviewed mostly performed CI/CT in San Bernardino.

**Supplementary Table 1-1**. Qualitative study participants characteristics

|  | n (%) | | |
| --- | --- | --- | --- |
| **Stakeholders:** | 24 (100) | | |
| Kaiser Permanente (KP) | 9 (38) | | |
| Public Health Institute (PHI) | 13 (54) | | |
| Other | 2 (8) | | |
|  | Total | Fresno | San Bernardino |
| **Micro-team (MT) members, n (row %):** | 23 (100) | 5 (22) | 18 (78) |
| Case investigator | 12 (52) | 3 (60) | 9 (50) |
| Resource coordinator | 4 (17) | 1 (20) | 3 (17) |
| Supervisor | 5 (22) | 1 (20) | 4 (22) |
| Team manager | 2 (9) | 0 (0) | 2 (11) |

Note: Column percentages are presented. Some participants were interviewed twice. Six MT members transferred their roles during the program period, but we reported the roles they maintained at the time of interview (for MT members still affiliated with the program) or the most recent roles (for MT members who left the program at the time of interview).
