## Supplementary Figure for "Completeness and timeliness of a contact tracing intervention during the COVID-19 pandemic: an outcome evaluation of the California Contact Tracing Support Initiative"

**Supplementary Figure 1.** Impact model for the CCTSI program. This diagram shows how program inputs (lavender) and case investigation and contact tracing (CI/CT) activities (pink) link to measurable program outputs (orange), which drive program health and employment outcomes (green) and impact (blue).

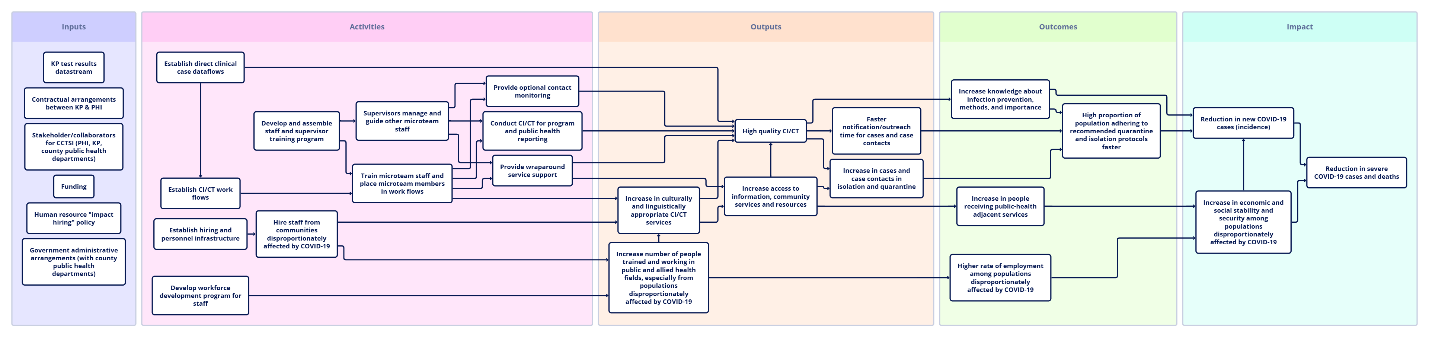

**Supplementary Figure 2**. Number of cases assigned, attempted, reached, and interviewed in Fresno County from December 2020 through July 2021

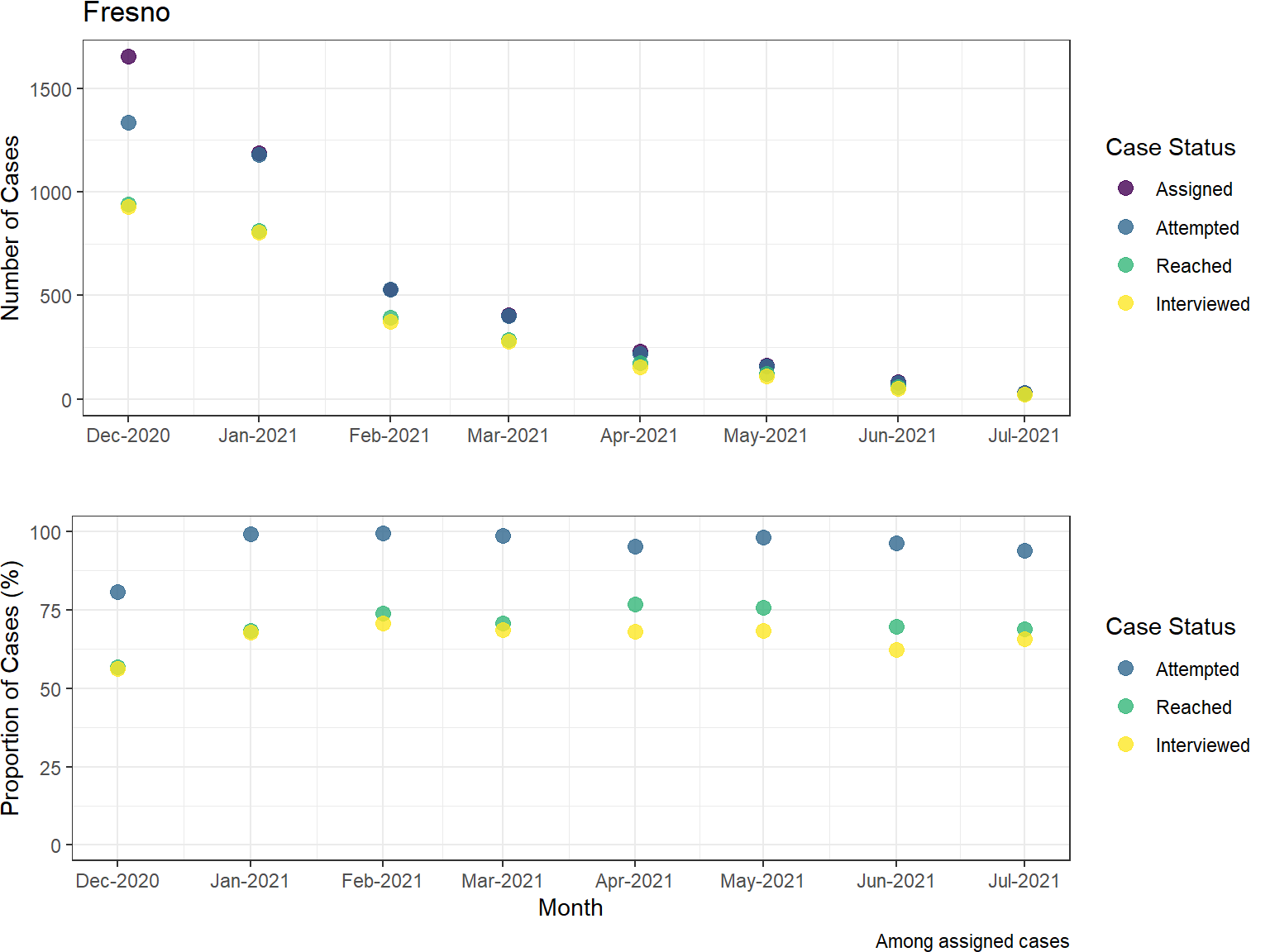

**Supplementary Figure 3**. Number of cases assigned, attempted, reached, and interviewed in San Bernardino County from December 2020 through July 2021

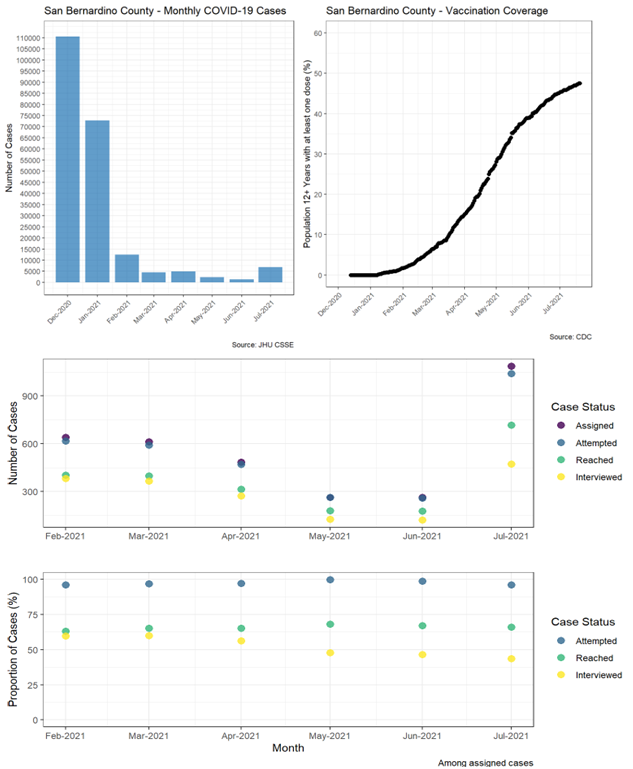

**Supplementary Figure 4**. Number of contacts assigned, attempted, reached, and notified in Fresno County from December 2020 through July 2021

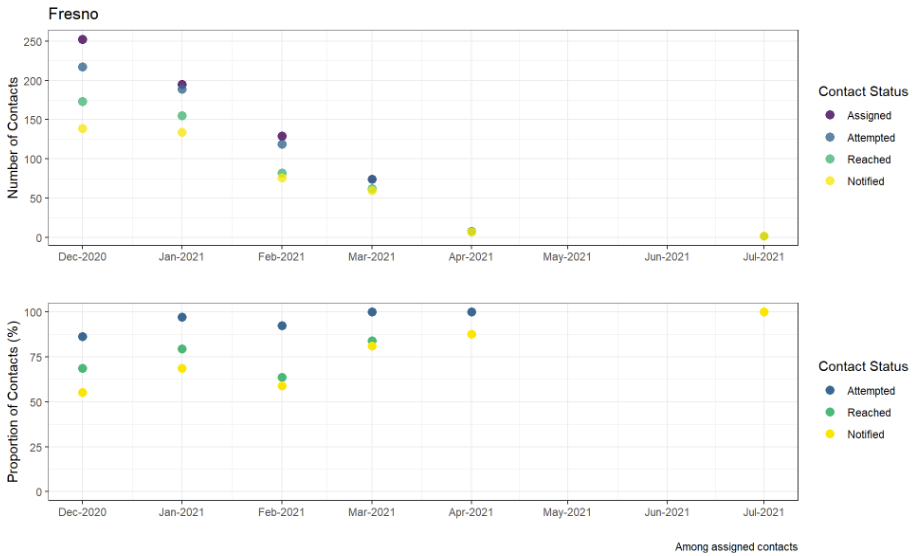

**Supplementary Figure 5**. Number of contacts assigned, attempted, reached, and notified in San Bernardino County from December 2020 through July 2021

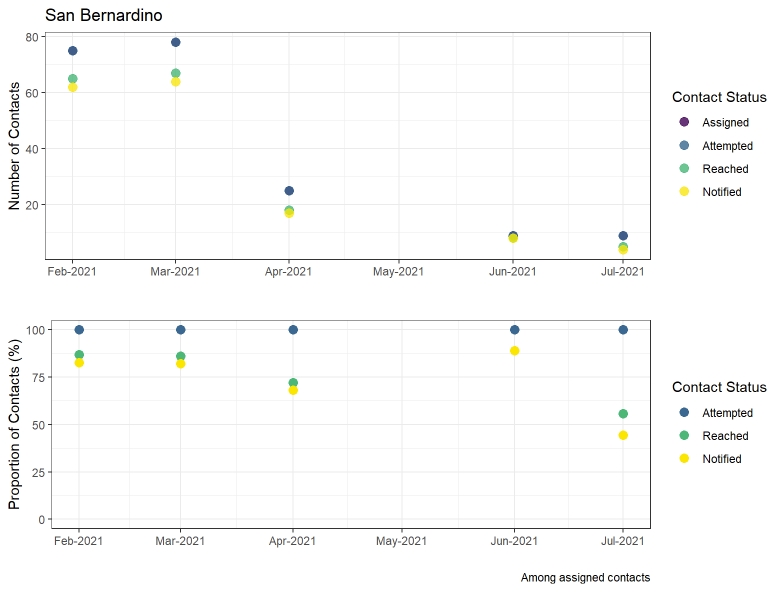

**Supplemental Table 1**. Cumulative completeness of case investigation and contact tracing (CI/CT) for the five county Local Health Jurisdictions (LHJs; randomly labeled as LHJ1-5)

|  | LHJ1 | LHJ2 | LHJ3 | LHJ4 | LHJ5 |
| --- | --- | --- | --- | --- | --- |
| Period of analysis | Jan 2021  – Jul 2021 | Dec 2020  – Jul 2021 | Dec 2020  – Jul 2021 | Dec 2020  – Jul 2021 | Dec 2020  – Jul 2021 |
| **Case investigations:** |  |  |  |  |  |
| Imported cases | N/A | N/A | N/A | N/A | N/A |
| Assigned | 36,874 | 9,174 | 7,612 | 1,687 | 168,933 |
| Attempted  (% of assigned) | 35,828 (97.2) | 8,839  (96.3) | 7,491  (98.4) | 1,664  (98.6) | 167,707 (99.3) |
| Reached  (% of assigned) | 23,996  (65.1) | 6,357  (71.9) | 6,512  (85.5) | 1,492  (88.4) | 148,478  (87.9) |
| Interviewed  (% of assigned) | 22,325 (60.5) | 6,234  (68.0) | 6,473  (85.0) | 978  (58.0) | 88,315 (52.3) |
| **Contact notifications:** |  |  |  |  |  |
| Total contacts elicited | 8,505 | 15,720 | 8,826 | 241 | 12,519 |
| Assigned (% of total contacts elicited) | 6,815  (80.1) | 3,146  (20.0) | 5,433  (61.6) | N/A | 6,717  (53.7) |
| Attempted  (% of assigned) | 6,043  (88.7) | 2,479  (78.8) | N/A | 237 | 6,148  (91.5) |
| Reached  (% of assigned) | N/A | 2,260  (71.8) | N/A | 222 | 5,314  (79.1) |
| Notified  (% of assigned) | 4,563  (67.0) | 1,511  (48.0) | N/A | 180 | 3,417  (50.9) |

Note: N/A indicates the information that the evaluation team did not receive from the LHJs.

**Supplemental Table 2**. Cumulative timeliness of case investigation and contact tracing (CI/CT) for the five county Local Health Jurisdictions (LHJs; randomly labeled as LHJ1-5)

|  | LHJ1 | LHJ2 | LHJ3 | LHJ4 | LHJ5 |
| --- | --- | --- | --- | --- | --- |
| Period of analysis | Jan 2021  – Jul 2021 | Dec 2020  – Jul 2021 | Dec 2020  – Jul 2021 | Dec 2020  – Jul 2021 | Dec 2020  – Jul 2021 |
| **Case investigations:** |  |  |  |  |  |
| Assigned cases | 36,874 | 9,174 | 7,612 | 1,687 | 168,933 |
| **From specimen collection to case import** |  |  |  |  |  |
| Within 1 day | N/A | N/A | N/A | N/A | N/A |
| Between 1-2 days | N/A | N/A | N/A | N/A | N/A |
| Average time (days) | 2 | 2.4 | 3.5 | 2.4 | 4.2 |
| **From case import to first outreach** |  |  |  |  |  |
| Within 1 day | 9,849 | 6,743 | 6,266 | 1,498 | 76,284 |
| Between 1-2 days | N/A | 2,431 | 545 | 80 | 22,987 |
| Average time (days) | N/A | N/A | N/A | 0.4 | 2.4 |
| **From case import to case interview** |  |  |  |  |  |
| Within 1 day | 4,575 | 3,825 | 5,517 | 697 | 37,037 |
| Between 1-2 days | 2,372 | 775 | 471 | 169 | 13,665 |
| Average time (days) | 6.8 | N/A | 2.2 | 1.5 | 9.0 |
| **Contact notifications:** |  |  |  |  |  |
| Assigned contacts | 6,815 | 3,146 | 5,433 | N/A | 6,717 |
| **From contact elicitation to first outreach** |  |  |  |  |  |
| Within 1 day | 4,865 | 1,499 | N/A | N/A | 3031 |
| Between 1-2 days | N/A | 149 | N/A | N/A | 213 |
| Average time (days) | N/A | N/A | N/A | N/A | N/A |
| **From contact elicitation to notification** |  |  |  |  |  |
| Within 1 day | 3,377 | N/A | N/A | N/A | 2,452 |
| Between 1-2 days | 268 | N/A | N/A | N/A | 172 |
| **From case specimen collection to contact notification, Average time (days)** | N/A | N/A | N/A | N/A | 2.3 |
| **From case data import to contact notification, Average time (days)** | N/A | 6.4 | N/A | N/A | 1.1 |

**Supplementary Table 3.** Wrap-around service (WAS) requests in two CCTSI sites (i.e., Fresno, San Bernardino)

|  | Fresno | San Bernardino |
| --- | --- | --- |
| Period of analysis | Dec 2020 – Jul 2021 | Feb 2021 – Jul 2021 |
| Total number of interviewed cases and notified contacts | 3,140 | 1,898 |
| WAS requests created, n (% among total interviewed and notified beneficiaries) | 1,342 (43) | 392 (21) |
| Requests closed, n (% of WAS request created) | 1256 (94) | 221 (56) |
| Services needed, n (% of WAS request created)^†^ |  |  |
| Food | 1073 (80) | 88 (22) |
| Housing | 139 (10) | 43 (11) |
| Mental Health | 0 (0) | 6 (1.5) |
| Utilities | 1074 (80) | 63 (16) |

^†^ Beneficiaries could request multiple services; thus, the percentage of services needed does not sum up to 100.

Note: For county data, only information relevant to cases/contacts needing isolation/quarantine support were available in LHJs 1 (n=30, 0% of interviewed cases and notified contacts), 2 (n=155, 2% of interviewed cases and notified contacts), and 5 (n=1,584, 2% of interviewed cases and notified contacts). We did not have information on whether beneficiaries were reached out for wrap-around services. LHJs 3 and 4 did not provide any information on wrap-around services.
